# Aligning computational pathology with clinical practice for colorectal cancer

**DOI:** 10.1101/2025.06.10.25329328

**Authors:** Elias Baumann, José F Carreño-Martinez, Ana Leni Frei, Javier Garcia-Baroja, Mauro Gwerder, Amjad Khan, Rina Mehmeti, Jacob Hanimann, Philipp Zens, Heather Dawson, Alessandro Lugli, Inti Zlobec

## Abstract

Pathology reporting of colorectal cancer (CRC) follows the International Collaboration on Cancer Reporting (ICCR) guidelines which define a set of 25 diagnostic report elements. To further develop the CRC diagnostic routine, multiple computational tools have been proposed in the last years. Despite the excellent sensitivity and potential advantages, many tools do not reach clinical deployment, suggesting that there are critical challenges to address when developing these algorithms. To summarize existing efforts in deep learning for ICCR CRC elements and highlight existing gaps between development and clinical deployment, this systematic review collected studies on computational tools for colorectal cancer histopathology analysis published between 2015 and 2024. Most of the 66 included studies focus on a subset of just three ICCR elements, namely mismatch repair status, BRAFV600E mutation testing, and lymph node status. Moreover, many of the studies did not include clinically relevant and validated results. These results show the gap between research and clinical practice in pathology with the example of CRC diagnosis. There is an unmet need for publicly available datasets, and a stronger focus on clinically important tasks. This review will contribute to aligning computation pathology with the clinic to increase the translational potential of developed tools.

## 1. Introduction

Digital pathology is shaping the future of the fast-evolving field of pathology and digitalized pathology laboratories are an increasing trend worldwide^1^. Once these laboratories have the digital infrastructure in place (e.g. slide scanner and viewer, laboratory information systems or image management system etc.), they can benefit from AI-based tools such as AI algorithms in supporting pathologists to analyze tissue slides. Leveraging AI is potentially helpful for time-consuming cases that require assessment of long lists of criteria and are prone to interobserver variability. Moreover, several studies have shown increased diagnosis confidence among pathologists when assisted with AI^2–4^.

AI-based tools have been developed in numerous different pathologies, including colorectal cancer (CRC). CRC is the third most common cancer worldwide with a raising incidence in younger ages due to sedentary lifestyle and consumption of processed food ^5^. As a result of this, and widespread screening, CRC constitutes a considerable portion of cases in a pathologist’s diagnostic routine.

On one hand, the World Health Organization (WHO) Classification covers all generally accepted criteria^6^ for standardized disease definition and classification. On the other hand, the College of American Pathologists (CAP) and ICCR have developed evidence-based guidelines for standardizing reporting in pathology that are implemented globally^7,8^. In this review, we focused on the ICCR CRC guideline^7^ which covers both the macroscopic as well as microscopic evaluation of CRC resection specimen. As automatic methods have been predominantly developed for microscopic assessment, we restrict the review to only include reporting elements from this category. These elements are classified into “core” elements that are key for disease management, staging and prognosis, such as histological tumor type and grade, lymphatic invasion, TNM staging; and “non-core” elements, recommended but not validated by all levels of evidence, including tumor budding, mismatch repair (MMR) status, coexistent pathology, etc. While various AI algorithms have been developed to assess individual core and non-core elements, it remains unclear whether they can be combined for an AI-based evaluation of all elements on the digitized whole slide image (WSI) and produce an automated CRC pathology report. With this in mind, through this systematic review we aimed to reflect on:

1. The current landscape of deep learning (DL) in computational pathology (CPath) for CRC
2. Published work on the 25 elements of the ICCR CRC diagnostic report
3. Gaps towards automated ICCR-based reporting
4. The potential avenues and future steps to close these gaps

## 2. Methods

### 2.1 Structure of this review

Given the breadth of this review, the Results section is divided into three major parts, starting with general meta-results on gathered papers, and followed by meta-analysis of each element of the ICCR CRC guideline grouped here as core or non-core. The elements are sorted by number of included publications. After this analysis, and a general discussion on the most important takeaways, we include recommendations for elements where currently no readily usable computational approach has been published (Fig. 1).

**Figure 1:**
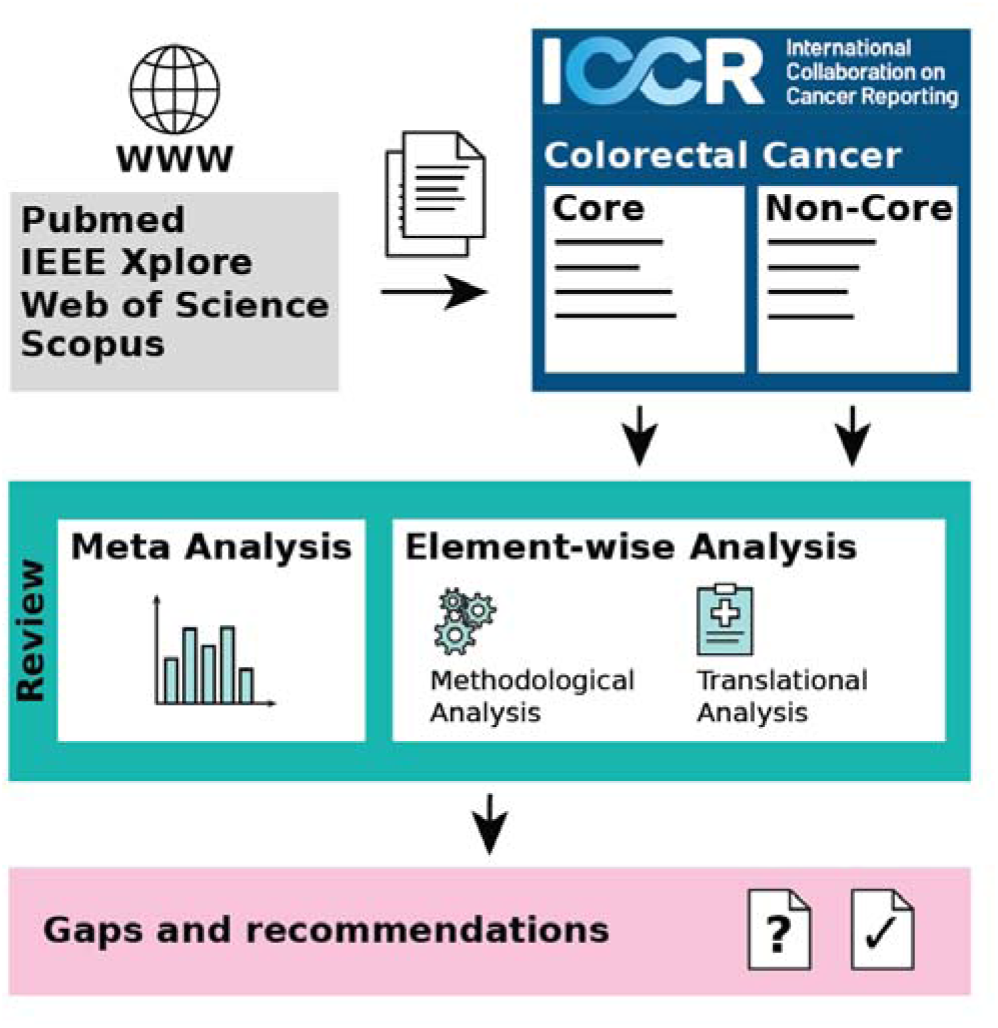
Graphical overview of the review.

#### Review Protocol

We perform a systematic literature review by following the Preferred Reporting Items for Systematic Reviews and Meta-Analysis (PRISMA) guidelines^9^.

#### Search strategy

For the review, we defined the following search criteria, with slight adaptations specific to the search engine. Essentially, search terms included a term related to cancer, to colon/colorectal, and a term related to histopathology to focus on the specific disease. Furthermore, deep learning, artificial intelligence, computer vision, and computer assistance-related search terms were added to cover all papers that develop computer-aided approaches. To include a wide range of publications across disciplines in our study, we included Pubmed, Web of Science, Embase, and IEEE Xplore as databases. Only papers published between January 1^st^ 2015 and September 13^th^ 2024 were eligible for inclusion in this study. Only abstracts in English language were included. All article abstracts were screened in covidence^10^. The full search terms for each database can be found in Supplementary Table 1.

#### Article Screening and eligibility

Articles were screened by at least two raters, with a third rater reviewing the abstract in case of disagreement. Duplicates were removed automatically by covidence^10^, or manually, if the automatic method failed to recognize the duplicate. Initially, abstracts were screened to assess relevance for the review according to a list of exclusion criteria. Specifically, articles should include an evaluation on human CRC, and results on histopathology image data. Articles that did not include original research (such as reviews, letters and commentaries) as well as conference abstracts were also excluded. Moreover, multimodal studies were excluded if they did not include results using exclusively histopathology/clinical data. If the approach evaluated non-hematoxylin and eosin (H&E) images, did not report slide-level or patient-level results, or only worked in a semi-automatic fashion they were also excluded. Finally, the article had to be related to or directly address one of the elements, e.g. if a study concerned itself with tissue type segmentation but is not usable for or applicable to one of the elements, it was excluded. Moreover, the developed model had to produce an output closely related to how the respective element is specified in the ICCR CRC guideline. In a second round, full texts are assessed for eligibility. The same exclusion criteria were applied. The full list of exclusion criteria can be found in Supplementary Table 2.

#### Adding references from other sources

Additionally, some articles from other sources highly relevant to this review were included. These papers were identified via references or by using search engines such as google scholar or semantic scholar for the specific ICCR CRC guideline element. Moreover, some articles did not contain the necessary information of the search criteria in their abstract, but were still relevant to this review, so they were manually included as well.

#### Data extraction

To thoroughly assess individual studies, information was extracted from each study. Specifically, data concerning the cohorts used (including staining, number of patients, cancer stage distribution, target distribution), the training setup (dataset splits, cross validation and involvement of pathologists), the DL pipeline (input size, pre-processing, model, post-processing, training task and target) and reported results (AUC, F1, … if available). Additionally, we assessed for each study whether there were slide-level or patient-level results, if the published model is clinically approved, whether data, code, or model weights were available and whether there is any information on inference time. Data were extracted by a single reviewer and verified by a second reviewer. Many studies report multiple results across several cohorts and in multiple configurations. To properly represent those studies in our analysis, the best results were extracted from each paper unless specified otherwise in the specific chapter. For multimodal models which use data that would normally not be used in a pathology report, we report the results where only histopathology data is used. Papers which cover more than one ICCR element will be included multiple times in the analysis but evaluated specifically for the respective topic.

## 3. Results

The search across databases yielded 4863 papers in total, of which 1636 were identified as duplicates. Additionally, 20 papers from citation search or additional manual search were included, yielding 3247 studies to be screened. In accordance with the exclusion criteria, 3066 studies were subsequently excluded, leaving 181 studies for full text screening. After full text screening, 66 studies were included in the final review (Figure 2).

**Figure 2:**
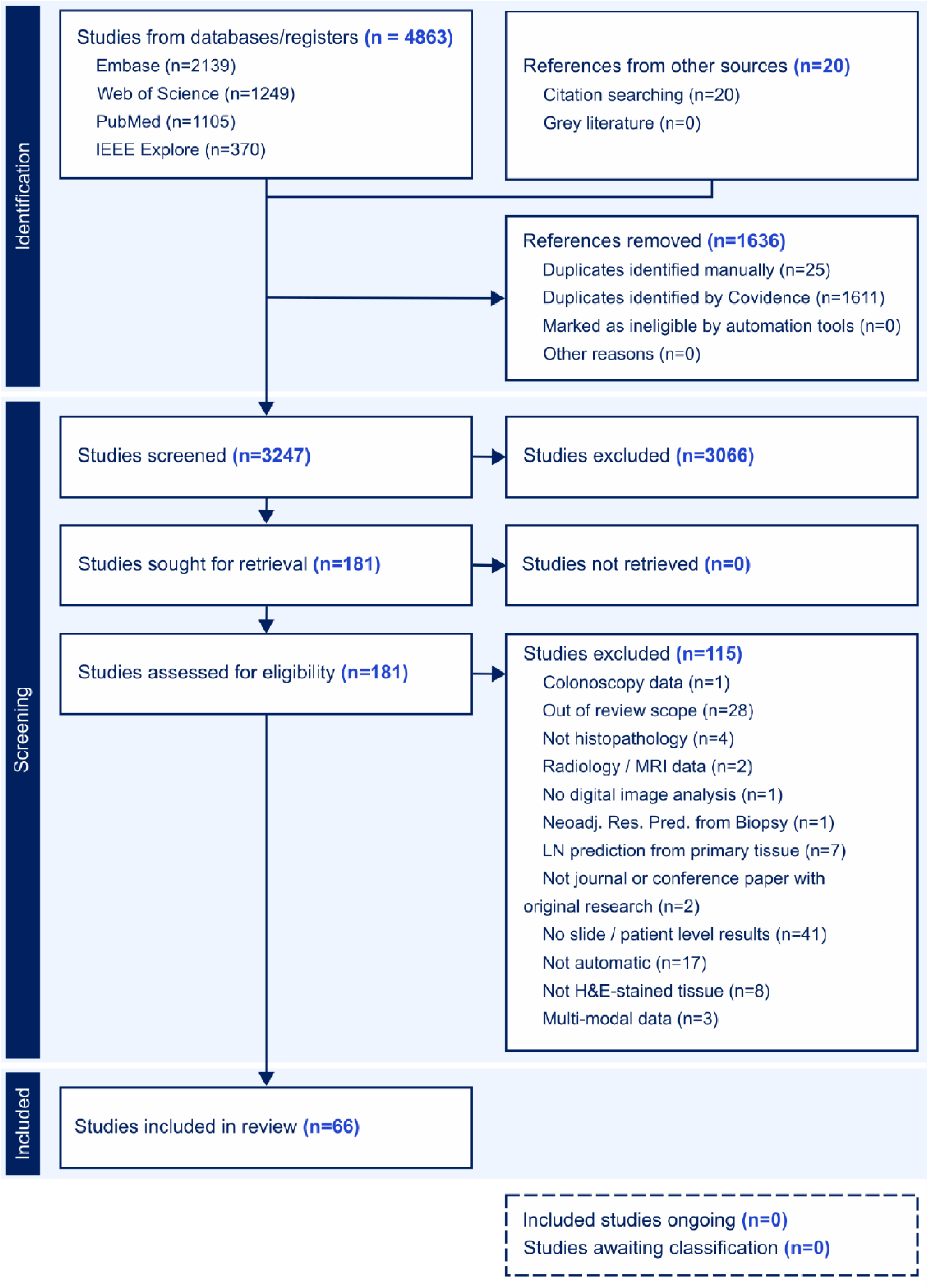
Flowchart following the PRISMA guidelines.

### 3.1 Meta Results

Although half of the ICCR CRC report elements are covered by the 66 selected studies, most publications concern only three topics. Specifically, 39/66 (59%) of studies showed prediction results on MMR status, 16/66 (24%) on BRAF V600E mutation, and 9/66 on pN (14%). Coexistent pathologies (5/66), Grade (4/66), and TNM Stage (3/66) are covered by five or less publications. For tumor budding and perineural invasion two studies were included each, whereas one study each for pT and response to neoadjuvant therapy was extracted. Publications can cover multiple topics at once, for example, 14 publications cover both MMR status and BRAF V600E mutation. For the remaining elements of the ICCR CRC guidelines the employed search and exclusion criteria did not yield any publications (Figure 3 A).

**Figure 3:**
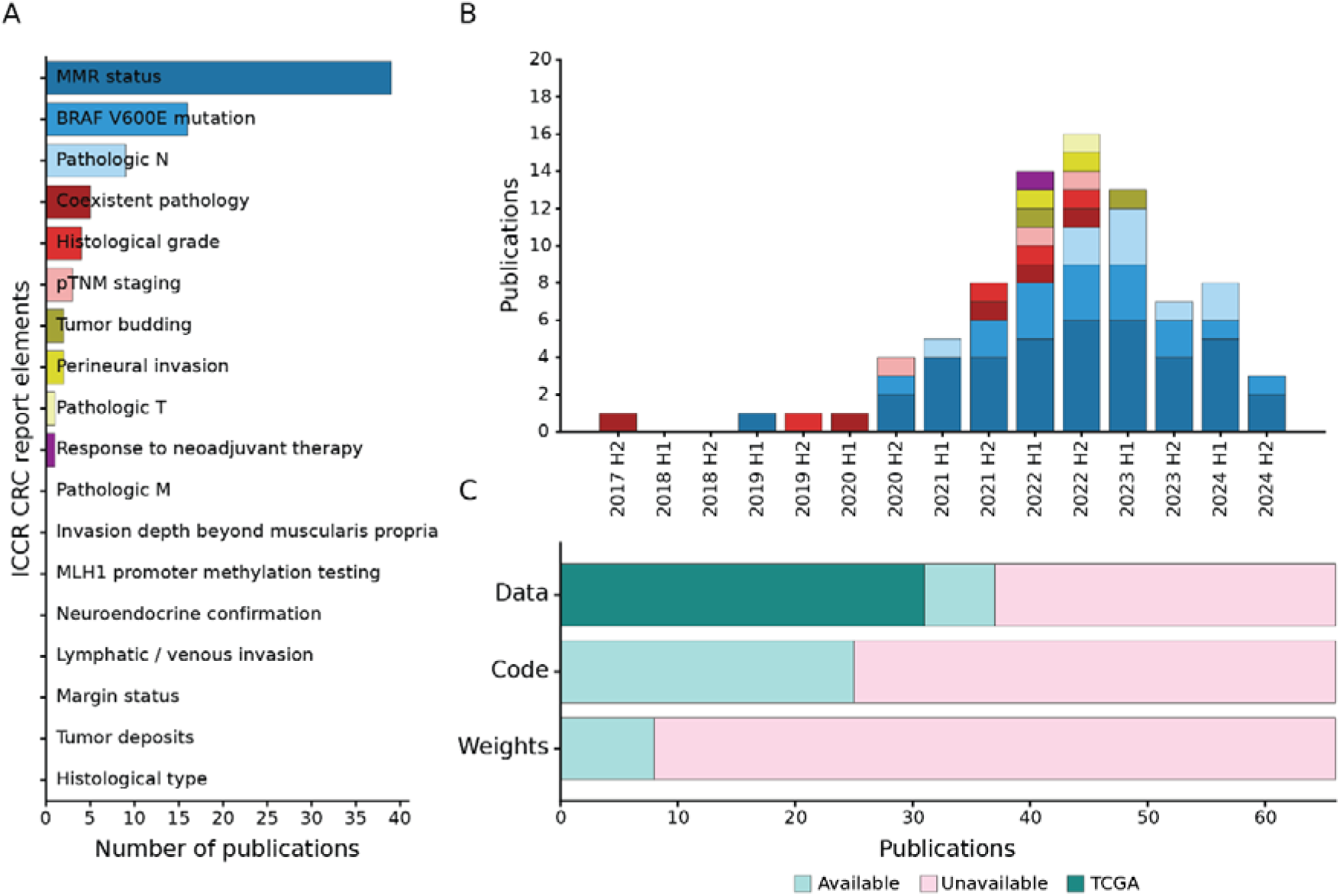
A) Number of articles covering a specific element of the ICCR guidelines. B) Number of published articles over time, aggregated by half-year. Studies for the second half of 2024 were only retrieved until 13.09.2024. C) Number of papers with accessible data, code and model weights respectively.

Following the general trend of increased attention and impactful contributions in CPath^11^, we also observe an overall increase in publications over time, even when applying strict exclusion criteria (Figure 2 B). MSI and BRAF mutation prediction studies remain the most common over the last four years. Lymph node metastasis prediction studies were increasingly published starting in the second half of 2022. Overall, the topic variety among published studies also increased in the past five years, but without a substantial growth in the number of publications in popular topics since 2022.

Considering data, code and model weight availability (Figure 2 C), more than half of the published papers (37/66, 56%) use some publicly available dataset or make their data available, and 31/66 (47%) use The Cancer Genome Atlas (TCGA) in some form. Code was available for less publications (25/66, 38%) and publications that made model weights available were scarce (8/66, 12%). It should be noted that resources which are available “upon reasonable request” were considered unavailable, although we did not investigate whether the information could have indeed been obtained in these cases.

This initial analysis has shown substantial variation in coverage across all reporting elements. In the next step, we will analyze published work related to the core elements of the ICCR CRC guideline.

### 3.2 ICCR CRC Guidelines: Core Elements

Elements classified as “Core” within the ICCR CRC guidelines are considered essential for clinical management and staging of CRC patients. Excluding clinical and macroscopically retrieved information, the microscopic core elements comprise histological tumor type, tumor grade, extent of invasion (and pT), lymphatic, venous and perineural invasion, lymph node status (and pN), tumor deposits, response to neoadjuvant therapy, margin status, margin status, histologically confirmed distant metastasis (and pM), the aggregation into pTNM, and confirmatory evaluation for neuroendocrine neoplasms. Of these twelve core elements, six have been addressed in 20 published articles, with pN (9/20) and tumor grade (4/20) receiving the most attention. 5/20 publications have used publicly available data or released their datasets, 6/20 made their code available, but none published model weights (Fig. 4). Next, we present the state of research of each individual core element in order of the number of included publications.

**Figure 4:**
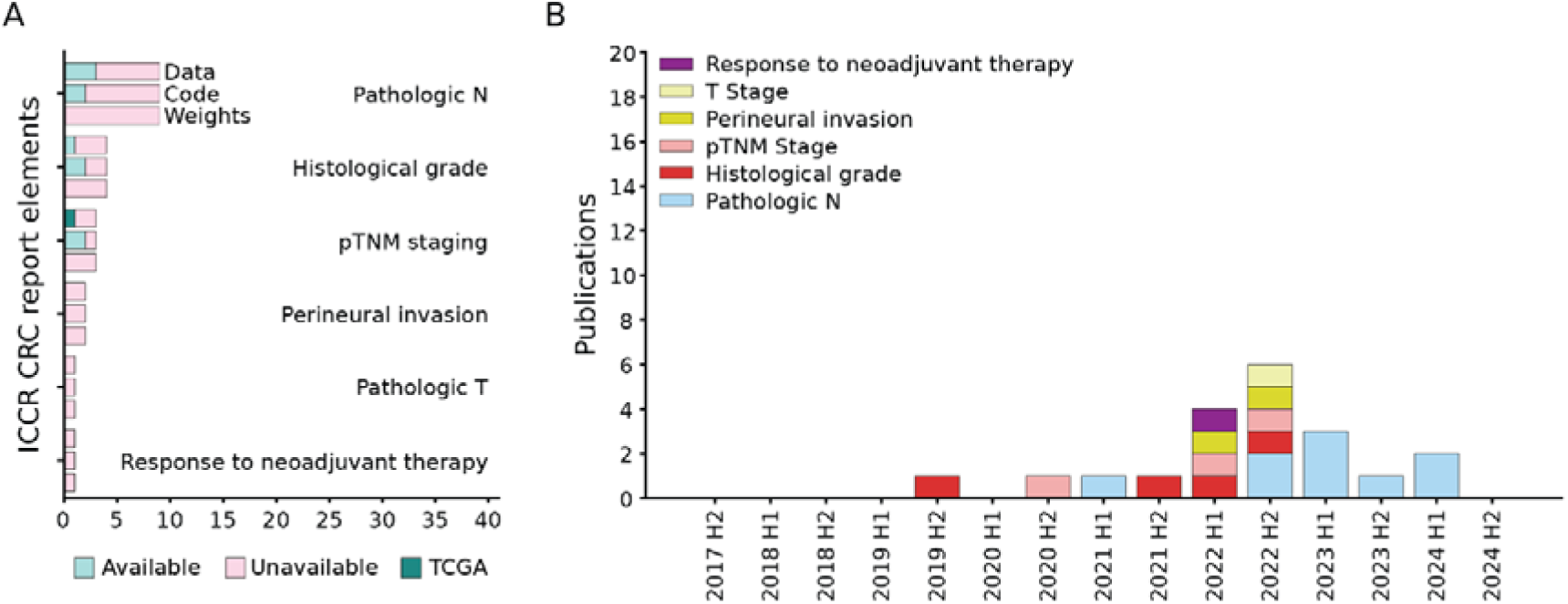
A) Number of articles covering a specific core element of the ICCR guidelines, split by the data, code, and weight availability for each topic. B) Number of published articles over time, aggregated by half-year. Not shown are elements where no publications passed exclusion criteria.

#### Regional Lymph Nodes (pN) and Lymph Node Status

Regional lymph node (LN) status is crucial for determining adjuvant chemotherapy and cancer staging. The TNM classification of regional LNs (pN) includes: NX (nodes cannot be assessed), N0 (no metastasis), N1a (1 positive node), N1b (2-3 positive nodes), N1c (tumor deposits in soft tissue without nodal metastasis), N2a (4–6 positive nodes), and N2b (≥7 positive nodes)^12^. A LN is considered positive if it contains at least one metastatic lesion larger than 2 mm^12^. Pathologists aim to evaluate at least twelve nodes per case, though this number can be influenced by factors such as specimen length, patient age, or neoadjuvant therapy^7,13^.

##### Data and Methods

Several approaches have been used to automatically screen LN slides for metastases, listed in Table 1. Most methods are focused on metastasis detection/segmentation or based on metastasis detection further predicting the slide-level labels, whether positive or negative. Model evaluation was performed on both external (30% of the studies) and internal cohorts (70% of the studies). Clinical validation or pathologist involvement was reported in ∼30% of studies.

**Table 1:**
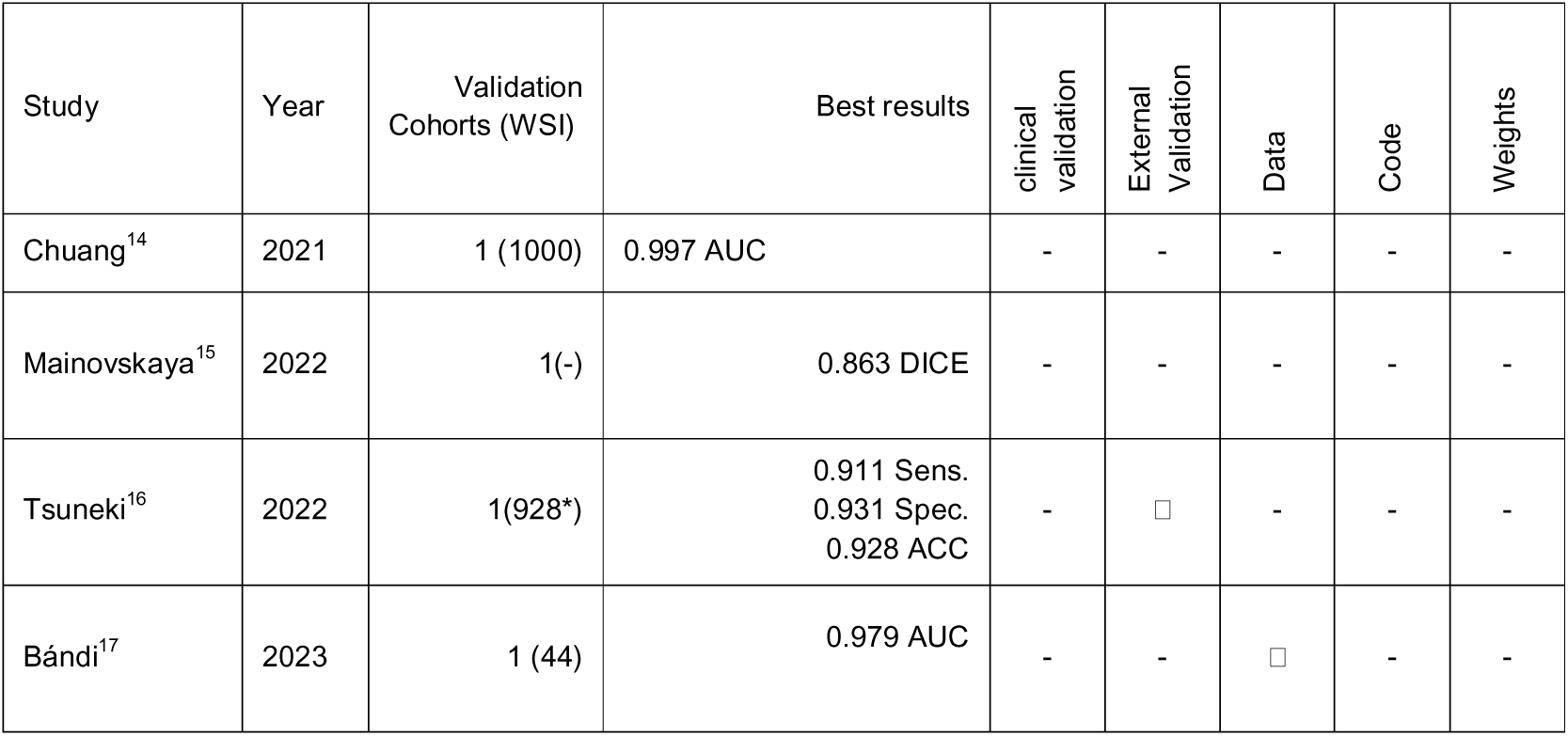
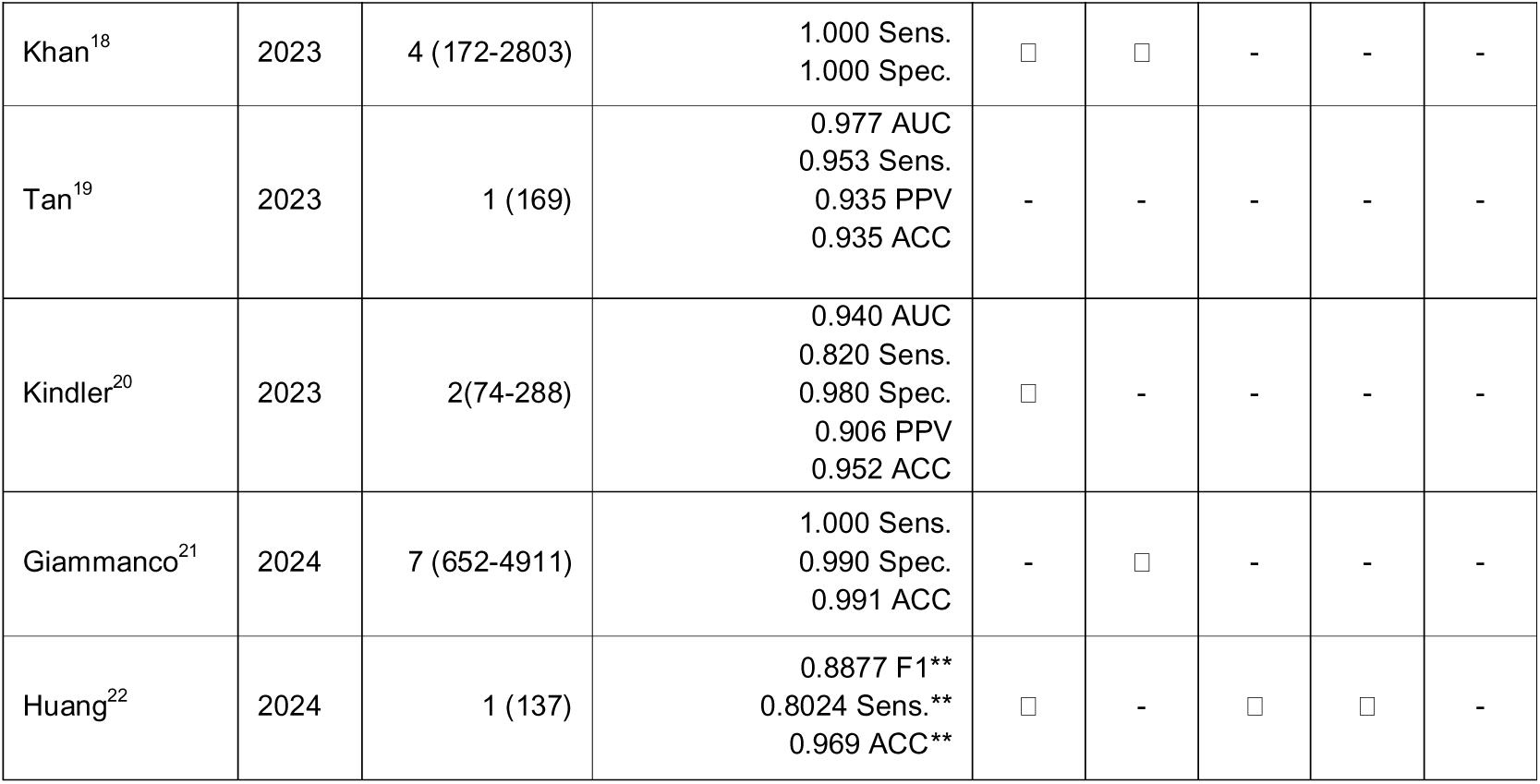
Extracted studies for lymph node status and pN. *187 CRC WSI, ** Pathologist performance assisted by model.

| Study | Year | Validation Cohorts (WSI) | Best results | clinical validation | External Validation | Data | Code | Weights |
| --- | --- | --- | --- | --- | --- | --- | --- | --- |
| Chuang<sup>14</sup> | 2021 | 1 (1000) | 0.997 AUC | - | - | - | - | - |
| Mainovskaya<sup>15</sup> | 2022 | 1(-) | 0.863 DICE | - | - | - | - | - |
| Tsuneki<sup>16</sup> | 2022 | 1(928*) | 0.911 Sens.<br>0.931 Spec.<br>0.928 ACC | - | □ | - | - | - |
| Bándi<sup>17</sup> | 2023 | 1 (44) | 0.979 AUC | - | - | □ | - | - |
| Khan<sup>18</sup> | 2023 | 4 (172-2803) | 1.000 Sens.<br>1.000 Spec. | □ | □ | - | - | - |
| Tan<sup>19</sup> | 2023 | 1 (169) | 0.977 AUC<br>0.953 Sens.<br>0.935 PPV<br>0.935 ACC | - | - | - | - | - |
| Kindler<sup>20</sup> | 2023 | 2(74-288) | 0.940 AUC<br>0.820 Sens.<br>0.980 Spec.<br>0.906 PPV<br>0.952 ACC | □ | - | - | - | - |
| Giammanco<sup>21</sup> | 2024 | 7 (652-4911) | 1.000 Sens.<br>0.990 Spec.<br>0.991 ACC | - | □ | - | - | - |
| Huang<sup>22</sup> | 2024 | 1 (137) | 0.8877 F1**<br>0.8024 Sens.**<br>0.969 ACC** | □ | - | □ | □ | - |

Huang et al.^22^ used a human-in-the-loop AI system (nuclei.io) to personalize ML models for colorectal LN metastasis detection. The approach improved accuracy, F1 score, sensitivity, and especially detection of isolated tumor cells, while significantly reducing evaluation time for fellows and negative cases. Similarly, Kindler et al.^20^ trained a Deep Neural Network tool achieving high pixel-level accuracy. Clinical testing showed high sensitivity (0.990), strong interobserver agreement (κ = 0.94), and significantly reduced review times for pathologists. Furthermore, Khan et al.^18^ demonstrated 100% agreement between their ensemble model and expert pathologists in a study of 217 cases.

##### Clinical Employability

The reviewed studies show high sensitivity and specificity across the board for metastasis detection. However, the automation of pN staging requires the additional step of counting the number of positive lymph nodes in a slide. This can be difficult due to tissue preparation steps where some large lymph nodes may be cut in half and may be present twice in the same slide, or on multiple different slides. Correctly re-identifying the same lymph node and ensuring positive lymph nodes not being counted twice therefore remains an unsolved problem. One approach to addressing this issue could be a better link between macroscopic tissue preparation and AI-based image analysis. For example, lymph node placement information could be additionally stored, but consistent tissue inking may also be an avenue to solve the problem automatically.

#### Histological Tumor Grade

Tumor grade describes the level of differentiation of tumor cells where a higher grade indicates a loss of glandular organization^6,23^. In accordance with the WHO classification, the ICCR CRC guidelines classify the grade by the least differentiated component of the neoplastic lesion which then results in either low or high tumor grade.

**Table 2:** Extracted studies for tumor grade. Normal (NORM), Well-differentiated (WD), Moderately-differentiated (MD), Poorly-differentiated (PD).

| Study | Year | Classes | Validation Cohorts (WSI) | Best Results | External Validation | Data | Code | Weights |
| --- | --- | --- | --- | --- | --- | --- | --- | --- |
| Rathore<sup>24</sup> | 2019 | WD, MD, PD | 1 (174) | 0.98 AUC<br>0.928 Sens.<br>0.958 Spec.<br>0.95 Acc | - | □ | - | - |
| Schrammen<sup>25</sup> | 2021 | NORM, WD+MD, PD | 1 (805) | 0.751 AUC<br>0.877 Sens.<br>0.528 Spec.<br>0.599 Acc<br>0.513 F1 | □ | - | □ | - |
| Chen<sup>26</sup> | 2022 | NORM, WD, MD, PD | 1 (108) | 0.981, 0.983, 0.963, 0.963 AUC | - | □ | - | - |
| Soldatov<sup>27</sup> | 2022 | NORM, WD, MD, PD | 1 (357) | 0.981, 1.0, 0.933, 1.0 Spec.<br>0.553, 0.0, 0.577, 0.0 Sens.<br>0.838, 0.871, 0.876, 0.974 Acc<br>0.696, 0.0, 0.6, 0.0 F1 | - | - | - | - |

##### Data and Methods

The retrieved studies use both private and public datasets, but with the exception of one study^25^, none validate their results externally. Given the subjectivity and high interobserver variabililty of tumor grade assessment^28^, a consensus label based on multiple pathologists might be more reliable as ground truth. Some studies^26,27^ used two raters, with a third pathologist to resolve any conflicts. Yet, none of the retrieved studies reported agreement between pathologists and deep learning models. Most studies rely on tile classification^25–27^ to directly predict tumor grade. Rathore et al.^24^ use morphological and texture-based image features aggregated in a support vector machine classifier and majority voting. They achieve comparable performance to tile classifiers on their dataset. For tile classification methods, the overall strategies include multiple instance learning^25^ for end-to-end training, and supervised tile-based training^26,27^. No studies aggregate at the patient level, although clinical practice typically considers the highest grade across multiple slides. This is particularly relevant for TCGA-based studies^26^, as WSIs were not selected for grading and may not be representative of the overall tumor grade.

##### Clinical Employability

Data accessibility varies significantly across studies. While Rathore et al.^24^ use the publicly available GLaS dataset and Chen et al.^26^ TCGA-COAD and TCGA-READ, and Soldatov et al.^27^ only make their dataset available upon request. Beyond TCGA and challenge datasets, independent validation is limited due to restricted access to additional clinical data.

Reproducibility is another concern, as most studies do not share code or model weights. Only Schrammen et al.^25^ have a publicly available repository. Despite these limitations, the availability of public datasets (e.g., GLaS) enables further research and tool development.

#### pTNM

Pathological TNM staging is the combined assessment of pT, pN, and pM. The TNM stage directly informs treatment decisions including whether a resection is necessary, or whether additional adjuvant therapy needs to be performed. It remains the most important prognostic factor in CRC diagnosis, with Stage I patients’ five-year survival being over 90% for colon and rectum, but Stage IV 11-15% respectively^29^.

**Table 3:** Extracted studies for pTNM. *Results from Image-only approach, paper reports improved results when integrating genomic data.

| Study | Year | Validation Cohorts (WSI) | Best Results | Actionable Results | External Validation | Data | Code | Weights |
| --- | --- | --- | --- | --- | --- | --- | --- | --- |
| Levy<sup>30</sup> | 2021 | 1 (17) | 0.92 AUC (Stage I,II vs III,IV) | - | - | - | □ | - |
| Kumar<sup>31</sup> | 2022 | 2 (130-197) | 0.78 - 0.81 AUC (Stage II vs IV) | □ | - | - | □ | - |
| Pei<sup>32</sup> | 2022 | 1 (459) | 0.89 Acc, 0.89 F1 (tile level) | - | - | □ | - | - |

While TNM Staging could be algorithmically constructed out of evaluated T, N, and M, all three retrieved approaches directly predict TNM stage from the primary, with only one publication also considering lymph node slides^30^. Two publications rely on TCGA COAD (and READ) for the evaluation, and Kumar et al.^31^ additionally include an internal cohort for training and validation. Two methods used graph convolution to spatially integrate tissue tile embeddings into slide level predictions^30,32^, whereas the last followed the more traditional method of tumor area detection, and subsequent tile classification and averaging based on morphological features of nuclei^31^.

##### Clinical Employability

Only Kumar et al.^31^ generate clinically actionable results, but they use their model for the specific task to differentiate between metastatic colon cancer and local colon cancer. While Pei et al.^30^ also show slide-level results, they never evaluate them or report a tile-aggregation strategy. Levy et al.^30^ predict low (I, II) vs high (III, IV) stage, but their approach does evaluate the slide for invasion depth so further development could lead to a comprehensive TNM analysis.

Taken together, none of the retrieved papers demonstrate an end-to-end pipeline that can reliably assess the TNM stage (I-IV) from histological slides. While there are interesting cases where directly predicting the risk of metastasis from the primary might be sensible (e.g. what is the risk of a missed metastasis), simpler approaches following the UICC TNM guidelines by aggregating information about local, regional and distant spread may be more clinically relevant for routine reporting.

#### Perineural Invasion

Perineural Invasion (PNI) is defined as tumor growth along a nerve, where the tumor surrounds at least one-third of the nerve’s perimeter and invades any of its layers^33^. PNI is reported as either present or absent, and its presence has been linked to poor prognosis, particularly in Stage II^34^.

**Table 4:** Extracted studies for perineural invasion.

| Study | Year | Validation cohort (WSI) | Best results | External validation | Data | Code | Weights |
| --- | --- | --- | --- | --- | --- | --- | --- |
| Han<sup>35</sup> | 2022 | 1 (50) | ACC: 0.81 | - | □ | - | - |
| Jung<sup>36</sup> | 2022 | 1 (-) | AUC: 0.92 | - | - | - | - |

##### Data and Methods

Both included studies employ a similar approach for detecting PNI. First, nerve and tumor regions are separately segmented, and subsequently a boundary between the two regions is identified. While Jung et al.^36^ use a private dataset for training, Han et al.^35^ trained on the PAIP2021 dataset comprising of 240 WSIs from multiple cancer types. The dataset was used for the PAIP2021 challenge^37^ to detect PNI.

##### Clinical Employability

Neither of the included studies provide the code or the weights of the DL algorithms that produced the reported results. Moreover, the validation cohorts, compared to studies on other topics, e.g. the previously discussed lymph node metastasis detection, are comparatively small. Beyond this reproducibility gap, the methods do report a binary output for the perineural invasion in accordance with the ICCR CRC guidelines.

#### pT and Extent of invasion

The extent of invasion of the primary tumor is the first part of the TNM staging and reflected in pT. It is assessed by considering the different components of the colon wall and the presence of tumor within them. pT ranges from pT0 (no tumor) to pT4 (tumor invades other organs or perforation into the visceral peritoneum) and has a direct impact on the treatment of a patient^12^.

**Table 5:**
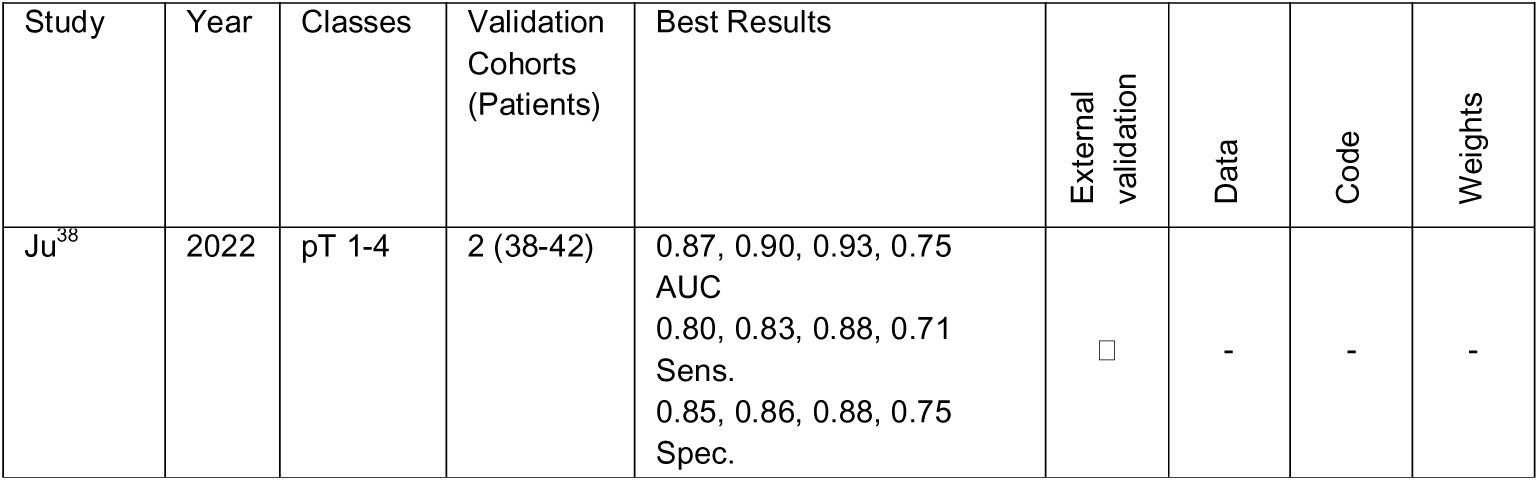
Extracted studies for extent of invasion and pT.

| Study | Year | Classes | Validation Cohorts (Patients) | Best Results | External validation | Data | Code | Weights |
| --- | --- | --- | --- | --- | --- | --- | --- | --- |
| Ju<sup>38</sup> | 2022 | pT 1-4 | 2 (38-42) | 0.87, 0.90, 0.93, 0.75<br>AUC<br>0.80, 0.83, 0.88, 0.71<br>Sens.<br>0.85, 0.86, 0.88, 0.75<br>Spec. | □ | - | - | - |

Only one study that automates the prediction of pT in CRC passed the exclusion criteria^38^. In this study, the authors use two validation cohorts, one internal with 38 patients, one external with 42 patients. For classification into pT1-4, they first segment the tumor area, then use a patch-based classifier to predict pT for each tile to finally choose the highest pT value as the final prediction. The model reaches AUCs up to 0.93 on the internal test set and 0.90 on the external validation cohort.

Yet the validation sets only include few patients, while data, code, and weights are not publicly available. They do show results with explicit cutoffs, but only in 1-vs-all settings without multiclass evaluation. Therefore, more work is needed to develop robust approaches for automatic pT assessment with larger validation cohorts and investigating challenges in real world datasets.

#### Response to Neoadjuvant Therapy

Tumor regression gradings (TRGs) are used to assess the response to neoadjuvant chemoradiotherapy (nCRT) by measuring the extent of tissue changes caused by the treatment^39^. This is a common assessment step in locally advanced rectal cancer treated with neoadjuvant therapies. Multiple TRGs exist but all rely on the assessment of tumor to fibrosis ratio within the tumor bed^39^.

**Table 6:** Extracted studies for response to neoadjuvant therapy.

| Study | Year | Classes | Validation cohort (Patients) | Best Results | External validation | Data | Code | Weights |
| --- | --- | --- | --- | --- | --- | --- | --- | --- |
| Zhang<sup>40</sup> | 2022 | Good response (TRG0-1), bad response (TRG2-3) | 1 (30) | 0.759 AUC<br>0.717 ACC | - | - | - | - |

##### Data and Methods

We retrieved only one study that predicted regression after nCRT in a fully automated way^40^. The authors used an MIL approach with gated attention weight normalization with a final bilinear attention for multi-scale feature fusion to classify responders from non-responders according to slide-level labels. Yet, they validated the model on a breast cancer metastasis dataset rather than rectal cancer which reduces the value of the validation.

##### Clinical Employability

There exist many different systems for TRG, and in the included study, the AJCC system^41^ was used. However, the four AJCC TRG categories were merged into a binary classification task which does not correspond to the ICCR CRC guidelines. The reviewed study trained for slide-level predictions but does not discuss aggregation strategies for patient-level predictions. However, this post-processing aggregation step is needed for successful clinical deployment, as pathologists report a patient-level TRG.

Due to the challenges caused by the TRG definitions, we did not find any study that presents a clinically ready algorithm to assess tumor regression in rectal cancer, yet a tissue type segmentation model fine-tuned to predict remaining tumor and fibrosis could replicate every TRG scheme.

### 3.3 ICCR CRC Guidelines: Non-Core Elements

The non-core elements of the ICCR CRC dataset assess additional features that provide valuable prognostic insights and are clinically relevant but are not frequently used in patient management or not yet validated. They include measurement beyond the muscularis propria, MLH1 promoter methylation, tumor budding, coexistent pathology, MMR status, and BRAF V600E mutation. Of these six elements, only four have been addressed in the published studies included in this review. MMR status and BRAF mutation, in particular, have received significant attention, with most MMR-related (28/39) and BRAF-related (11/16) studies using TCGA as their primary data source. For these elements, code to reproduce results is often available (21/48, 44%), and some studies provide access to trained model weights (8/48, 17%). Interestingly, since similar methods are applied for MMR– and BRAF-status prediction, 14 studies investigate both biomarkers simultaneously. In contrast, tumor budding and coexistent pathology remain comparatively unexplored and overall lack publicly available data, code, or access to weights (Fig. 5). In the following section, we examine the current research landscape for each of the non-core elements.

**Figure 5:**
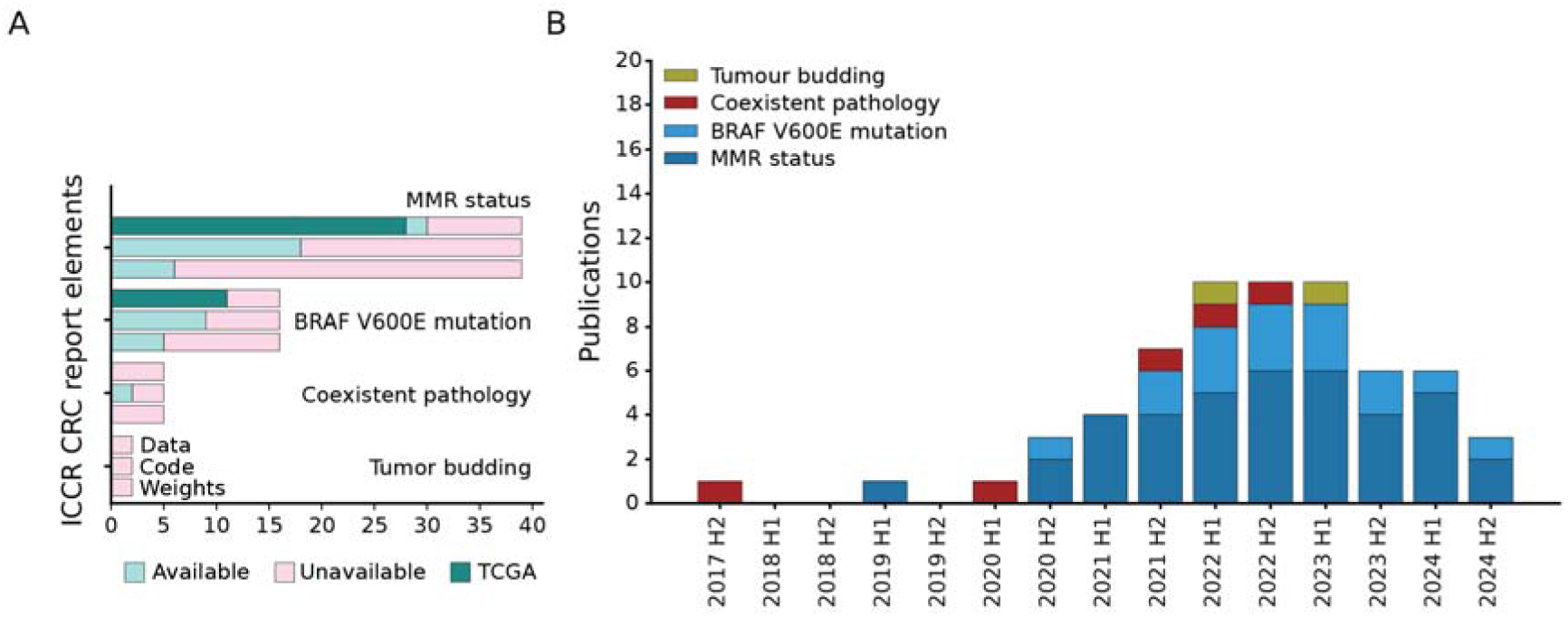
A) Number of articles covering a specific non-core element of the ICCR guidelines, split by the data, code, and weight availability for each topic. B) Number of published articles over time, aggregated by half-year. Not shown are elements where no publications passed exclusion criteria.

#### MMR status

The DNA mismatch repair (MMR) system is responsible to correct DNA replication mismatches^42^. The mutation of its major genes and the resulting instability of microsatellite repeat sequences is referred to Microsatellite Instability (MSI)^43^. MSI is a biomarker in CRC, guiding clinical decisions for intermediate risk Stage II and Stage IV CRCs^44,45^, and represents an indicator for lynch syndrome screening – a hereditary condition characterized by mismatch repair gene mutations. Routine diagnostic testing is typically performed using Immunohistochemistry (IHC) and both ASCO and ESMO guidelines mandate dMMR testing for all new CRC diagnoses ^46^. Recent DL models have demonstrated that they can predict MSI status directly from H&E slides without the need for IHC.

**Table 7:** Extracted studies for microsatellite instability. *Publication uses multimodal data to improve results, reported are pathology only, ** Results based on some form of training only on TCGA if available.

| Study | Year | Validation Cohorts (Patients) | Best AUC | TCGA COAD/READ (FFPE) AUC** | Actionable Results | External Validation | Data | Code | Weights |
| --- | --- | --- | --- | --- | --- | --- | --- | --- | --- |
| Kather<sup>47</sup> | 2019 | 3 (360-378) | 0.84 | 0.77 | - | □ | □ | □ | - |
| Schmauch<sup>48</sup> | 2020 | 1 (465) | 0.81 | 0.81 | - | - | □ | □ | - |
| Lee<sup>49</sup> | 2020 | 1 (57) | - | - | □ | - | □ | - | - |
| Yamashita<sup>50</sup> | 2021 | 3 (100-479) | 0.93 | 0.84 | □ | □ | □ | - | - |
| Ke<sup>51</sup> | 2021 | 1 (747) | - | - | - | - | □ | - | - |
| Pai<sup>52</sup> | 2021 | 2 (136-230) | 0.852 | - | □ | - | - | - | - |
| Lee<sup>53</sup> | 2021 | 3 (91-500) | 0.97 | 0.89 | □ | □ | □ | - | - |
| Schrammen<sup>54</sup> | 2021 | 2 (889-2448) | 0.91 | - | □ | □ | - | □ | - |
| Saillard<sup>55</sup> | 2021 | 3 (47-555) | 0.97 | 0.88 | - | □ | □ | - | - |
| Bilal<sup>56</sup> | 2021 | 3 (47-428) | 0.98 | 0.86 | - | □ | □ | - | - |
| Bustos<sup>57</sup> | 2021 | 1 (1788) | 0.9 | - | □ | - | - | - | - |
| Echle<sup>58</sup> | 2022 | 9 (51-2452) | 0.96 | 0.91 | □ | □ | □ | □ | □ |
| Saldanha<sup>59</sup> | 2022 | 5 (632-2448) | 0.89 | - | - | □ | □ | □ | - |
| Laleh<sup>60</sup> | 2022 | 2 (632-2448) | 0.93 | 0.89 | - | - | □ | □ | - |
| Fujii<sup>61</sup> | 2022 | 3 (248-986) | 0.92 | - | - | - | - | □ | □ |
| Jiang<sup>62</sup> | 2022 | 5 (78-441) | 0.88 | 0.88 | □ | □ | □ | - | - |
| Guo<sup>63</sup> | 2022 | 1 (396) | 0.76 | 0.76 | - | - | - | - | - |
| Lou<sup>64</sup> | 2022 | 3 (5-144) | 0.94 | - | □ | - | - | - | - |
| Teichmann<sup>65</sup> | 2022 | 1 (840) | 0.94 | - | - | - | □ | - | - |
| Lv<sup>66</sup> | 2022 | 1 (868) | 0.92 | 0.92 | - | - | □ | - | - |
| Rubinstein<sup>67</sup> | 2022 | 1 (313) | 0.71 | 0.71 | - | - | □ | - | - |
| Ding<sup>68</sup> | 2022 | 3 (94-306) | 0.84 | 0.84 | - | □ | □ | □ | - |
| Cao<sup>69</sup> | 2023 | 2 (47-360) | 0.96 | 0.91 | □ | □ | □ | - | - |
| Guo<sup>70</sup> | 2023 | 2 (423-1065) | 0.91 | 0.91 | □ | □ | - | - | - |
| Chang<sup>71</sup> | 2023 | 2 (305-1579) | 0.95 | 0.72 | □ | □ | □ | □ | - |
| Niehues<sup>72</sup> | 2023 | 2 (2190-2448) | 0.94 | - | □ | - | - | □ | □ |
| Yuan<sup>73</sup> | 2023 | 1(491) | 0.93 | - | - | - | □ | - | - |
| Tsai<sup>74</sup> | 2023 | 2 (628-927) | 0.88 | 0.88 | □ | □ | □ | □ | - |
| Wagner<sup>75</sup> | 2023 | 17 (35-2452) | 0.96 | 0.83 | □ | □ | □ | □ | - |
| Cen<sup>76</sup> | 2023 | 2 (425-1138) | 0.92 | 0.92 | - | - | □ | □ | - |
| Kim<sup>77</sup> | 2023 | 1 (118) | 0.92 | - | - | - | □ | - | - |
| Saillard<sup>78</sup> | 2023 | 4 (47-554) | 0.96 | 0.93 | □ | □ | □ | - | - |
| Zamanitajeddin<sup>79</sup> | 2024 | 2 (47-428) | 0.99 | 0.88 | - | □ | □ | - | - |
| Roth<sup>80</sup> | 2024 | 2 (110-632) | 0.89 | 0.89 | - | - | □ | □ | - |
| Liu<sup>81</sup> | 2024 | 1 (352) | 0.69* | - | - | - | - | - | - |
| Shen<sup>82</sup> | 2024 | 1 (100) | 0.85 | 0.85 | - | - | □ | - | - |
| Gustav<sup>83</sup> | 2024 | 4 (27-2039) | 0.94 | 0.87 | □ | □ | □ | □ | - |
| Vorontsov<sup>84</sup> | 2024 | 2 (360-482) | 0.96 | - | - | - | □ | □ | □ |
| Wang<sup>85</sup> | 2024 | 3 (75-437) | 0.88 | 0.87 | - | □ | □ | □ | □ |

##### Data and Methods

Common datasets for evaluation are the publicly available TCGA COAD/READ cohorts (80.4%), the PAIP2020 challenge dataset (26.3%), and DACHS (21.05%). Of note here is that different publications use different subsets of TCGA COAD/READ so while results have been generated on some of the same slides metrics cannot be properly compared. At the same time, many papers only used one (33.3%) or two (35.3%) cohorts in their analysis and the remainder used more (29.4%).

There are mixed approaches to ground truth, as for MSI/dMMR testing, several different methods are clinically appropriate. 33.3\% relied on both IHC and PCR for the same or different datasets, 49.0% only rely on PCR, 11.8% only on IHC, 3.9% on other methods. None of the papers using IHC as gold standard are reporting whether these were single observer results, even though there is some disagreement in manual IHC dMMR protein analysis^86^.

Most MSI prediction models rely on tile level embeddings that are aggregated to a slide-level score. Before that step, many approaches pre-detect the tumor area (43.6%), and then subsequently usually add an average (41.1%), majority (23.5%) or top-k (17.6%) aggregation on top. Models that did not pre-segment the tumor tissue (56.4%) commonly relied on an attention mechanism (50%) to weight tile results for the final slide-level MSI score.

Foundation models have included MSI prediction in CRC on datasets like TCGA-CRC-DX^47,87^ as a benchmark task ^80,84,85^, which will lead to more results in this topic, though the small scope of the evaluation may not bring additional insights on clinical translatability.

##### Clinical Employability

In MSI/dMMR prediction, the DL model either needs to predict MSI as well as a diagnostic test (e.g. at least as well as IHC) or needs to be used as a screening tool. In both cases, a cutoff needs to be defined. In this review, 47.1% of papers defined some cutoff at least on a subset of the validation data, but only 13.7% demonstrated some applicable cutoff viable for screening (e.g. at least 90% sensitivity) in a clinical setting. One method is clinically approved as a pre-screening tool with CE-IVD certification^78^, yet so far, no tool can reach the same specificity as IHC or PCR testing while maintaining 95% sensitivity (Saillard et al.^78^ 46% Spec. at 98% Sens., Wagner et al.^75^ 61% Spec. at 95% Sens.).

The current state of papers therefore indicates that the MSI category is advanced, yet tools can only be employed as screening tools with gold-standard testing necessarily following the screening as the specificity is comparatively low. Therefore, either DL models can be further improved to have similar sensitivity and specificity as gold-standard testing, or this step necessarily needs to be deferred to a non-H&E evaluation.

#### BRAF V600E

BRAF mutation testing plays an important role in Lynch syndrome identification after MSI testing. On the other hand, microsatellite stable BRAF mutant colorectal cancers are particularly aggressive, yet the mutation also increases responsiveness to EGFR inhibitor therapy^88^. The non-core element in the CRC ICCR guidelines specifically concerns the oncogenic V600E mutation, which represents approximately 90% of BRAF mutant cases in a large trial^89^. Notably, BRAF and MSI cases share similarities in both morphological features and epidemiological patterns^90^.

**Table 8:** Extracted studies for BRAF V600E mutation.

| Study | Year | Validation Cohorts (Patients) | Best AUC | TCGA COADREAD (FFPE) AUC | Actionable Results | External Validation | Data | Code | Weights |
| --- | --- | --- | --- | --- | --- | --- | --- | --- | --- |
| Fu<sup>91</sup> | 2020 | 1 (146) | 0.571 | 0.571 | - | - | □ | □ | □ |
| Bilal<sup>56</sup> | 2021 | 1 (502) | 0.79 | 0.79 | - | - | □ | - | - |
| Schrammen<sup>54</sup> | 2021 | 1 (2075) | 0.82 | - | □ | - | - | □ | - |
| Chen<sup>92</sup> | 2022 | 1 (414) | 0.729 | 0.729 | - | - | □ | - | - |
| Fujii<sup>61</sup> | 2022 | 3 (248-986) | 0.859 | - | □ | - | - | □ | □ |
| Guo<sup>63</sup> | 2022 | 1 (396) | 0.897 | 0.897 | - | - | - | - | - |
| Laleh<sup>93</sup> | 2022 | 2 (534-2081) | 0.86 | 0.808 | - | □ | □ | □ | - |
| Saldanha<sup>94</sup> | 2022 | 4(500-2075) | 0.77 | - | - | □ | □ | □ | - |
| Teichmann<sup>65</sup> | 2022 | 1 (268) | 0.75 | 0.75 | - | - | □ | - | - |
| Cen<sup>76</sup> | 2023 | 2 (500-1026) | 0.78 | 0.78 | - | □ | □ | □ | - |
| Guo<sup>70</sup> | 2023 | 2 (500-1026) | 0.8 | 0.77 | □ | □ | - | - | - |
| Niehues<sup>72</sup> | 2023 | 2 (1478-2081) | 0.82 | - | - | □ | - | □ | □ |
| Tsai<sup>74</sup> | 2023 | 2 (591-906) | 0.71 | 0.71 | □ | □ | □ | □ | - |
| Wagner<sup>75</sup> | 2023 | 9 (107-2081) | 0.88 | 0.78 | - | □ | □ | □ | - |
| Zamanitajeddin<sup>79</sup> | 2023 | 1 (502) | 0.79 | 0.79 | - | - | □ | - | - |
| Wang<sup>85</sup> | 2024 | 2 (233-559) | 0.732 | 0.732 | - | □ | □ | □ | □ |

##### Data and Methods

As there is considerable overlap between the setup, task, and patients with MSI and BRAF mutation, publications frequently predict both. Therefore, different subsets of the TCGA cohort are the also most prevalent datasets (75%), followed by the DACHS cohort (31.3%).

Most approaches apply stain normalization (68.8%) and tile extraction (100%) as preprocessing steps. Many workflows (31.3%) subsequently identify tumor tissue using a tissue classification algorithm^56,65,70,76,79^. The majority of publications implement MIL-based approaches (81.3%), differing in their tile aggregation approaches (averaging: 23.1%, top k tiles: 30.7%, other: 30.9%). These MIL-based approaches are commonly built on either CNN-based (31.3%) or attention-based (31.3%) architectures. Recent publications are showcasing foundation models (18.8%) with either averaging-based ^91,92^ or attention-based^85^ aggregation strategies.

##### Clinical Employability

Only 43.8% of publications conducted an external validation of their algorithm, while 56.3% utilized more than one cohort. Openly available datasets were included in 68.8% of all studies. The code is available in 62% of all publications, and model weights are only available in 31.25% of studies. Furthermore, most studies did not differentiate between BRAF^V600E^ mutations and all other BRAF mutations (87.5%). In contrast, Fuji et al. utilized a BRAF^V600E^-specific cohort, whereas Tsai et al. reported both BRAF^V600E^-specific and unspecific results.

Only four studies reported actionable results^54,61,70,74^, meaning that a label (BRAF^WT^/BRAF^mut^) rather than a mutation probability was assigned to each WSI. Only two studies report prediction sensitivities higher than 90% ^54^. Of those, only one study reached an accuracy higher than 0.5 ^54^.

Of all 16 included publications, only Guo et al.^70^ and Schrammen et al.^25^ report fully integrated pipelines with results based on a clinically appropriate cutoff (sensitivity >90%). However, Guo et al.^70^ neither shared code nor weights, and Schrammen et al.^25^ did not use any openly available datasets on which results could be reproduced.

The current state of BRAF^V600E^ prediction is unsolved and mostly serves as a validation task for multi-purpose models. Almost all (88%) studies reporting BRAF^mut^ –status report MSI-status as well. As those features are highly correlated, it is expected that models will perform similarly for both tasks. However, there exist cases that are non-correlating, which are especially interesting to pathologists^95^. None of the studies investigated performance in such cases. Furthermore, most studies do not utilize a clinically appropriate cutoff, making it hard to judge their performance in a clinical setting.

#### Coexistent Pathology

Under coexistent pathology, all other identified lesions should be listed. This could be additional synchronous carcinomas, polyps or e.g. IBD, other dysplastic normal tissue, subsumed under other lesions. Published work usually focuses on precursor lesions or the correct classification of polyps, but those models could be applied to e.g. correctly classify additional polyps in the same case. Additionally, because of the diversity of possible findings within this section, the models that have been developed so far only focus on one of the items that should be reported.

**Table 9:**
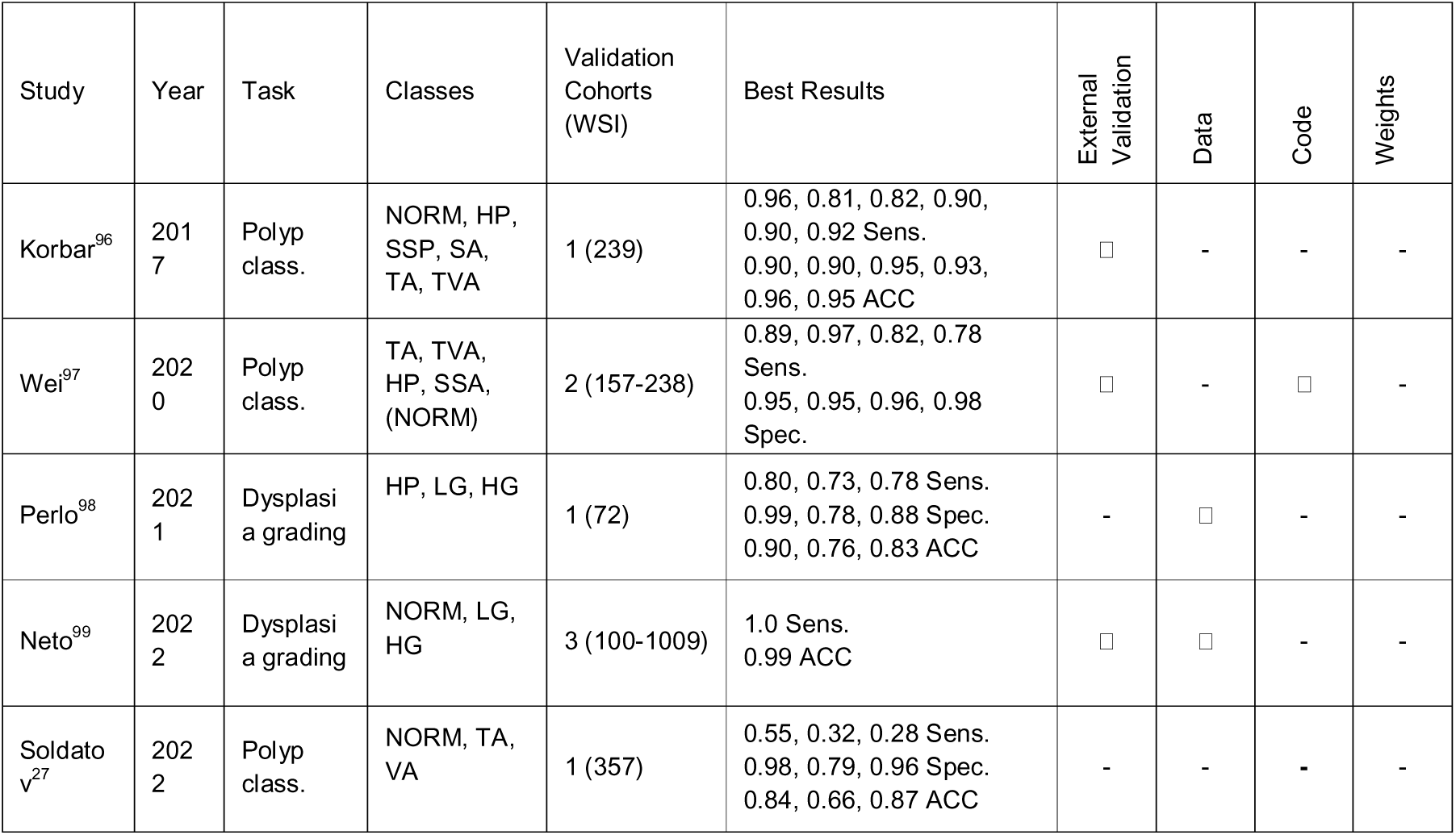
Extracted studies for response to coexistant pathology. NORM, Normal tissue; HP, Hyperplastic Polyp; TA, Tubular Adenoma; TVA, Tubulo-Villous Adenoma; SSP, Sessile Serrated Polyp; SA, Serrated Adenoma; SSA, Sessile Serrated Adenoma; LG, low grade dysplasia; HG, high grade dysplasia,

##### Data and Methods

The reviewed studies utilized datasets derived from biopsy or polyp resections rather than oncologic resection specimens. Across studies, there is lack of access to the development datasets.

It is important to highlight that the same task has different GT across studies. While only two studies address dysplasia grading, they incorporate different classes. Perlo et al.^98^ add Hyperplastic Polyp (HP) to their dysplasia categories, which is not used by Neto et al.^99^. Similar observations apply for polyp classification.

Slide-level classification is achieved by Multiple Instance Learning by Neto et al.^99^, while Perlo et al.^98^ aggregated tile classification results from a CNN. Wei et al.^97^ defined class-specific thresholds to categorize whole slide images (WSIs).

##### Clinical Employability

Variability in ground truth definitions limits model comparability and clinical integration. Differences in annotation standards hinder consistent evaluation, while a lack of publicly available datasets and independent validation cohorts restricts reproducibility. Only Neto et al.^99^ provide accessible validation data, but broader external validation remains scarce. Existing studies focus on polypectomy specimens rather than oncologic resection samples, since the studies are designed for application to polyp classification in the colorectal cancer screening workflow. While existing models can assess isolated polyps, their performance on primary tissue slides, which encompass all layers of the colon, and thereby unseen tissue types, is uncertain.

Furthermore, several coexisting pathologies remain unexplored. For instance, no studies were identified that automatically evaluate (IBD) using H&E-stained slides. While Rymarczyk et al.^100^ present a model to score Crohn’s disease and ulcerative colitis, it relies on prior classification into one of these two categories, highlighting a gap to achieve automatic reporting of this section of the guidelines.

#### Tumor Budding

Tumor budding (TB) is defined as presence of small clusters of up to four cancer cells at the tumor invasive front in a tissue section^101^. TB assessment in cancer follows the standardized guidelines of the International Tumor Budding Consensus Conference (ITBCC) and is performed in hotspot regions (0.785 mm²) of H&E-stained tissue sections^102^. This scoring has clinical implications, particularly for pT1 and stage II CRC patients, as high TB is associated with poor prognosis ^101,103^. Identifying and counting TBs on H&E slides is time-consuming and prone to inter-observer variability making it an ideal task to be automated by DL models.

##### Data and Methods

Most published studies on AI-assisted TB assessment rely on IHC staining, patch-level, single-center training and validation of the model. After exclusion criteria, two studies following the standardized ITBCC assessment pipeline were left (Table 10).

**Table 10:** Extracted studies for tumor budding.

| Study | Year | Validation cohorts (WSI) | Best Results | External validation | Tumor bud detection and count | Data | Code | Weights |
| --- | --- | --- | --- | --- | --- | --- | --- | --- |
| Lu<sup>104</sup> | 2022 | 1 (40) | 0.96 AUC<br>0.94 Sens.<br>0.83 Spec.<br>0.86 PPV | - | □ | - | - | - |
| Bokhorst<sup>105</sup> | 2023 | 2 (15-18) | 0.98 Sens.<br>0.44 PPV<br>0.58 F1 | □ | □ | - | - | - |

The study by Lu, et al.^104^ uses faster RCNN ^106^ for bud detection. Ground truth was established manually on H&E tissue slides and the model’s evaluation time was significantly faster than manual assessment.

On the other hand, Bokhorst et al.^105^ created a pipeline integrating a U-Net^107^ model for tissue segmentation and HoVer-Net^108^ for nuclei detection at the invasive front. The tumor bulk and invasive front were identified using a convex-hull algorithm. Their strategy was validated on data from four different medical centers.

##### Clinical Employability

Both models present different approaches in TB detection and counting in H&E WSIs that seem promising to reduce time and inter-observer variability among pathologists. However, there are a number of limitations to be addressed before being able to deploy such models in the diagnostic routine, such as small validation datasets, an overestimation of TB count especially in necrotic areas, and an inability to distinguish pseudo budding ^109^.

## 4. Discussion

In this large systematic review, we have analyzed published work on deep– and machine learning for digital pathology image analysis by putting each study into context of the ICCR CRC guidelines. Particular focus was put on whether studies could be implemented in a clinical workflow to assist pathologists by automating the CRC pathology report. This work has shown substantial gaps and numerous elements of the ICCR CRC guidelines which have not been addressed by any study.

Moreover, studies often do not publish code or weights such that they could be reproduced or implemented in an experimental workflow, and data is rarely shared as well.

One of the main observations from our analysis is that despite the critical role of core elements of the ICCR CRC guidelines for clinical decision making, most research efforts focus on automating the reporting of non-core elements. This discrepancy highlights the challenge in aligning the research objectives of the CPath community with the practical needs of the clinic. Many core tasks are not simple, and some are not investigated at all. Instead, they are often overlooked due to a perceived lack of methodological novelty, despite being central to real-world diagnostic workflows.

In this context, it is important to acknowledge that the recent advances in AI have powered the development of chatbots^110^, or computer vision tools that can work as an end-to-end diagnostic system or even predict patient survival^111^. While these models offer the appeal of simplified outputs by bypassing intermediate diagnostic variables, they risk overlooking the interpretability and clinical validation that the elements of the ICCR guidelines offer. In contrast, building trustworthy and reliable models for ICCR-defined elements aligns more closely with current diagnostic practice. Such an approach may facilitate adoption by pathologists and laboratories, as more transparent model outputs can be verified and causally understood^112^. On the other hand, in established diagnostic criteria, it can take decades to alter established criteria or add additional ones, such that end-to-end prognostic models may not have a chance to ever be adopted as standard-of-care.

Another pressing issue is the generalized lack of multi-cohort evaluation. Models are often developed and validated on single-institution datasets or within closed consortia, which limits generalizability. Differences in staining protocols, scanning equipment, and population demographics are known to significantly affect performance, and are a source of bias^113,114^, yet multi-site evaluation that would better reflect real-world conditions is rarely done, mostly due to data access barriers. As a result, many studies use TCGA data, but H&E slides in TCGA were never selected for diagnostic evaluation. Therefore, validation results on TCGA have only limited transferability to other real-world cohorts. This review excluded studies which could not report slide level results, but frequently research groups do not have access to clinical data or whole slide annotations to verify their results. Efforts such as BIGPICTURE ^115^ could help facilitate more clinically relevant research by making such data available.

Challenges have been crucial in pushing the boundaries of deep learning research in computational pathology^37,116,117^. Even after challenges have been completed, a number of studies still use the provided benchmark for further evaluation of new approaches. Yet, these challenges and benchmarks focus on a specific problem which may not be translatable to a diagnostic element and often do not require methods to develop deployable tools but rather highly specific methods to beat the benchmark. Moreover, inference time, economic considerations and required infrastructure are usually not addressed either, such that these challenges often are only useful for deep learning researchers, but not for clinicians and laboratory staff. On a positive note, challenges provide a standardized evaluation dataset such that methods can be compared, whereas even validation on TCGA is often done non-transparently without publishing the used data split, list of included slides in train and test set, or the exclusion criteria.

This review focused on the microscopic evaluation of CRC resection specimen and exclusively on H&E based evaluation, yet a diagnostic report would also contain macroscopic information or use other data modalities. Macroscopic analysis includes taking photos of the specimen and textual information exists, so theoretically, deep learning models could be developed for this purpose, too. Deep learning-based analysis of IHC, while more expensive since additional stainings are necessary, may be more reproducible and would still be automatically assessed^118^. Novel machines analyzing tissue before embedding could also further pave the way for additional deep learning tools improving diagnostic routine by creating different imaging data^119.^

Regardless, the major results of this review are the identification of several gaps in the ICCR criteria that current research has not yet covered. Table 11 gives an overview over all topics not yet comprehensively addressed by current work. For each topic, a potential approach is outlined, together with the required data and the potential benefit from automatically analyzing this element.

**Table 11:**
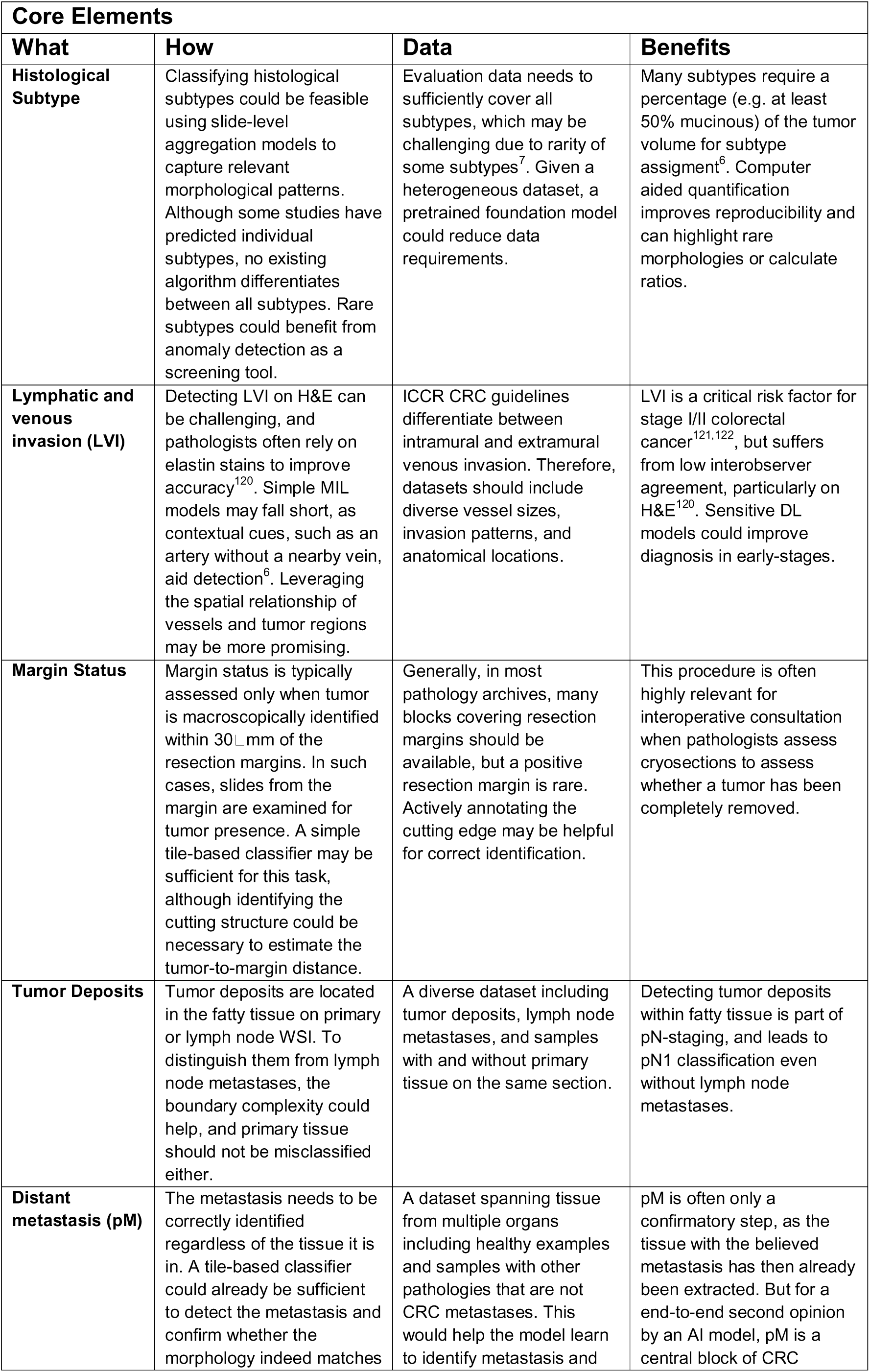
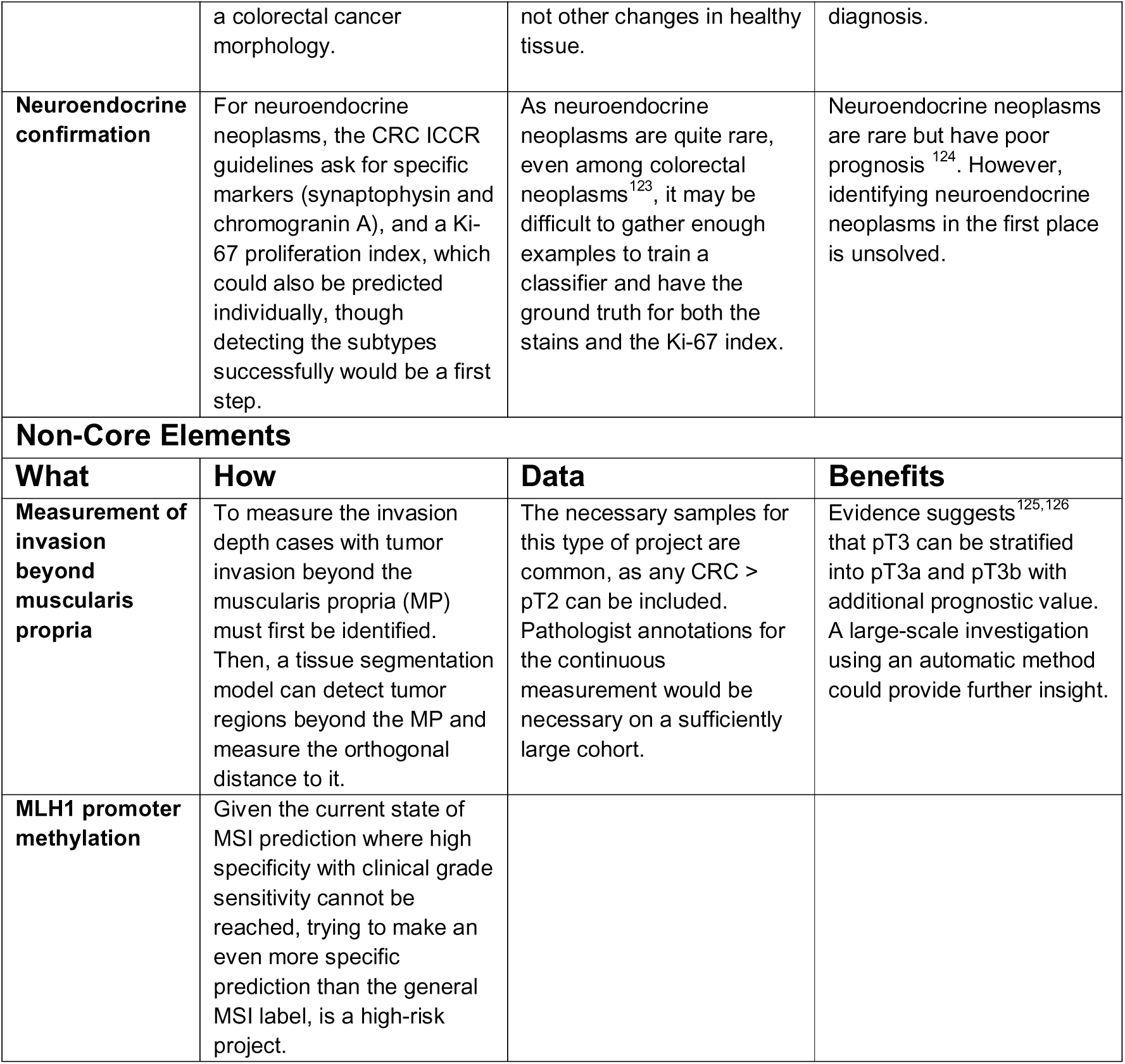
Addressing the remaining ICCR CRC guideline elements.

This overview could be a starting point for a number of investigations, yet it is important to mention that each element may have its own set of specific problems that only arise once researchers investigate the element.

## 5. Limitations

Limiting the scope of the review to papers with slide-level results and automated analysis ignores a sizable number of papers that could likely be translated to working WSI level approaches with little additional work. Yet, these excluded articles are also not less important for the progress in the field. Moreover, the employed search terms potentially excluded relevant papers if they did not include the indicated terms in the abstract. By selecting ICCR CRC guidelines over alternatives like CAP, we emphasized certain topics while omitting emerging biomarkers such as tumor-stroma ratio, tumor border configuration, and immune response. Additionally, the focus on H&E-based algorithms ignores immunohistochemistry-based approaches, which represent promising alternatives in CRC diagnostics.

## 6. Strengths

This systematic review includes publications on deep– and machine-learning AI-models for image analysis in pathology according to elements of the ICCR CRC dataset^7^ – a globally recognized, evidence-based and standardized pathology reporting guideline. It provides the readers with the current landscape of research in CPath towards automating routine tasks of pathologists during CRC assessment according to ICCR guideline. In the results section we provide a general analysis on the trend of publications over the years and per each element of the guideline. We also present information on the data, code and weight availability among included publications.

Following the general overview, we dissect publications individually, summarize their key results in separate tables as well as critically evaluate the clinical readiness of the proposed models, for each core and non-core element of the ICCR CRC guideline.

Moreover, to guide future research efforts, we present a dedicated table containing all underexplored core and non-core elements, where we propose a training approach, the dataset types and the anticipated benefits of utilizing such models.

Finally, we emphasize practical aspects for researchers, such as the necessity for transparency in computational requirements, availability of code, weights and data – essential for reproducibility and improvement of AI tools with clinical applicability.

All in all, this review should facilitate alignment between future research efforts in CPath and clinical practices as well as encourage new studies to fill current gaps and underexplored research areas.

## 7. Conclusion

This systematic review provides a comprehensive overview of the current state of CPath research to address elements of the diagnostic report in CRC. It reveals the limited maturity of the field and lack of alignment between research and clinical practices and emphasizes the need for developing approaches for various gaps while evaluating existing methods in clinical contexts. The findings could also lead to new benchmarks for novel models that assess entire diagnostic workflows rather than isolated tasks. Finally, this review underscores the remarkable complexity of CRC diagnosis performed by pathologists, which represents just one of many diseases they routinely evaluate every day.

## Data Availability

All data produced in the present study are available upon reasonable request to the authors.

## 8. Acknowledgements

This work was made possible via funding from several sources. E.B. and A.L.F. were funded by the Swiss National Science Foundation (CRSII5_193832). A.K. was funded by the Center for Artificial Intelligence in Medicine (CAIM), University of Bern. M.G. and J.F.C. was funded by the Swiss cancer league (KFS-5786-02-2023-R). J.H. was funded by the Institute of Tissue Medicine and Pathology, University of Bern. R.M. was funded by Swiss National Science Foundation (31003A_166578/1) and the Swiss Government Excellence Scholarship (ESKAS, nr. 2021.0019 / Kosovo / OP). J.G.B. was funded by the Swiss National Science Foundation (10.000.619).

We would also like to thank Tanya Karrer, University Library of Bern, for her valuable help in the conceptual development of this review.

