## Supplementary Material for "Aligning computational pathology with clinical practice for colorectal cancer"

**I.1 Individual Search Strategies**

| Database | Search Terms |
| --- | --- |
| Pubmed | (Cancer*[tiab] OR Tumo*r*[tiab] OR carcinoma*[tiab] OR neoplasm*[tiab] OR malignan*[tiab] OR "Neoplasms"[MeSH Terms]) AND (Colon[tiab] OR "colon"[MeSH Terms] OR Colorectal[tiab] OR Rectal[tiab] OR Rectum[tiab] OR "rectum"[MeSH Terms] OR Bowel[tiab] OR Colonic[tiab]) AND ("artificial intelligence"[tiab] OR "ai based"[tiab] OR "deep learning"[tiab] OR "Artificial Intelligence"[MeSH Terms] OR transformer[tiab] OR "convolutional neural network"[tiab] OR CNN[tiab] OR "machine learning"[tiab] OR "neural network"[tiab] OR "computer assisted"[tiab] OR "computer aided"[tiab] OR "computer vision"[tiab] OR "digital image analysis"[tiab] OR "image based"[tiab] OR "computational pathology"[tiab]) AND (patholog*[tiab] OR histopatholog*[tiab] OR histolog*[tiab] OR "Pathology"[MeSH Terms] OR "Histology"[MeSH Terms]) AND ("2015/01/01"[Date - Publication] : "3000"[Date - Publication]) AND English[Language] |
| Web of Science | (((((((TS=((Cancer* OR Tumor* OR Tumour* OR carcinoma* OR neoplasm* OR malignan*))) AND TS=((Colon OR Colorectal OR Rectal OR Rectum OR Bowel OR Colonic))) AND TS=(("artificial intelligence" OR "ai based" OR ai-based OR "deep learning" OR deep-learning OR transformer OR "convolutional neural network" OR CNN OR machine-learning OR "machine learning" OR "neural network" OR computer-assisted OR "computer assisted" OR computer-aided OR "computer aided" OR computer-assisted OR "computer assisted" OR "computer vision" OR "digital image analysis" OR image-based OR "image based" OR "computational pathology"))) AND TS=((patholog* OR histopatholog* OR histolog* )))) AND PY=(2015-2030)) AND LA=(English)) AND DT=(Article OR Early Access OR Proceedings Paper) |
| Embase | (cancer*:ti,ab,kw OR tumo?r*:ti,ab,kw OR carcinoma*:ti,ab,kw OR neoplasm*:ti,ab,kw OR malignan*:ti,ab,kw OR 'neoplasm'/exp) AND (colon:ti,ab,kw OR colorectal:ti,ab,kw OR rectal:ti,ab,kw OR rectum:ti,ab,kw OR bowel:ti,ab,kw OR colonic:ti,ab,kw OR 'colon'/exp OR 'rectum'/exp) AND ('artificial intelligence':ti,ab,kw OR 'ai-based':ti,ab,kw OR 'deep learning':ti,ab,kw OR transformer:ti,ab,kw OR 'convolutional neural network':ti,ab,kw OR cnn:ti,ab,kw OR 'machine learning':ti,ab,kw OR 'neural network':ti,ab,kw OR 'computer aided':ti,ab,kw OR 'computer assisted':ti,ab,kw OR 'computer vision':ti,ab,kw OR 'digital image analysis':ti,ab,kw OR 'image based':ti,ab,kw OR 'computational pathology':ti,ab,kw OR 'artificial intelligence'/exp OR 'machine learning'/exp OR 'convolutional neural network' OR 'computer vision'/exp) AND (patholog*:ti,ab,kw OR histopatholog*:ti,ab,kw OR histolog*:ti,ab,kw OR 'pathology'/exp OR 'histology'/exp) AND english:la AND [2015-2024]/py AND ([article]/lim OR [article in press]/lim OR [conference paper]/lim OR [preprint]/lim) AND [humans]/lim |
| IEEE Xplore | ("All Metadata":Cancer OR "All Metadata":Cancers OR "All Metadata":Tumo*r* OR "All Metadata":carcinoma OR "All Metadata":carcinomas OR "All Metadata":neoplasm* OR "All Metadata":malignan* OR "Mesh_Terms":"Neoplasms") AND ("All Metadata":Colon OR "All Metadata":Colorectal OR "All Metadata":Rectal OR "All Metadata":Rectum OR "All Metadata":Bowel OR "All Metadata":Colonic OR "Mesh_Terms":colon OR "Mesh_Terms":rectum) AND ("All Metadata":"artificial intelligence" OR "All Metadata":"ai based" OR "All Metadata":"deep learning" OR "All Metadata":transformer OR "All Metadata":"convolutional neural network" OR "All Metadata":CNN OR "All Metadata":"machine learning" OR "All Metadata":"neural network" OR "All Metadata":"computer aided" OR "All Metadata":"computer assisted" OR "All Metadata":"computer aided" OR "All Metadata":"computer assisted" OR "All Metadata":"computer vision" OR "All Metadata":"digital image analysis" OR "All Metadata":"image based" "All Metadata":"computational pathology" OR "Mesh_Terms":"Artificial Intelligence") AND ("All Metadata":patholog* OR "All Metadata":histopatholog* OR "All Metadata":histolog* OR "Mesh_Terms":"Pathology" OR "Mesh_Terms":"Histology") |

**Supplementary Table I.1.1:** Search criteria for each of the search engines. Included terms are the same, but some search tools support mesh terms or already include tags such as only conference and journal papers.

**I.2 Exclusion Criteria**

- Not histopathology (E.g. Colonoscopy imaging data, or Radiology data)
- No digital image analysis
- Not colorectal cancer
- Not English
- Not human tissue
- Not relevant to ICCR guidelines
- Not journal or conference paper with original research
- Retracted article
- Multimodal, if no unimodal pathology results are available
- Not automatic (needs manual input)
- No slide level / patient level results
- Not H&E

**Supplementary Table I.2.1:** Exclusion criteria applied for abstract screening and full text screening. If possible, they were already included in the search terms, otherwise articles were filtered at the screening stage.
